# ACE gene variants rise the risk of severe COVID-19 in patients with hypertension, dyslipidemia or diabetes. A pilot study

**DOI:** 10.1101/2021.03.24.21253576

**Authors:** María Íñiguez, Patricia Pérez-Matute, Pablo Villoslada-Blanco, Emma Recio-Fernandez, Diana Ezquerro-Pérez, Jorge Alba, M. Lourdes Ferreira-Laso, José A. Oteo

## Abstract

Coronavirus disease 19 (COVID-19) caused by the severe acute respiratory syndrome coronavirus 2 (SARS-CoV-2) infection continues to scale and threaten human health and public safety. It is essential to identify those risk factors that lead to a poor prognosis of the disease. A predisposing host genetic background could be one of these factors that explain the interindividual variability to COVID-19 severity. Thus, we have studied whether the rs4341 and rs4343 polymorphisms of the angiotensin converting enzyme (ACE) gene, key regulator of the renin-aldosterone-angiotensin system (RAAS), could explain the different outcomes of 128 COVID-19 patients with diverse degree of severity (33 asymptomatic or mildly asymptomatic, 66 hospitalized in the general ward, and 29 admitted to the ICU). We found that G allele of rs4341 and rs4343 was associated with severe COVID-19 in hypertensive patients, independently of gender (*p*<0.05). G-carrier genotypes of both polymorphisms were also associated with higher mortality (*p*< 0.05) and higher severity of COVID-19 in dyslipidemic (*p*<0.05) and type 2 diabetic patients (*p*< 0.01).

In conclusion, our preliminary study suggests that the G-containing genotypes of rs4341 and rs4343 confer an additional risk of adverse COVID-19 prognosis. Thus, rs4341 and rs4343 polymorphisms of ACE could be predictive markers of severity of COVID-19 in those patients with hypertension, dyslipidemia or diabetes. The knowledge of these genetic data could contribute to precision management of SARS-CoV-2 infected patients when admitted to hospital.

## INTRODUCTION

At the time of writing this paper, the COVID-19 has resulted in a pandemic with more than 100 million of confirmed cases around the world that has caused more than 2 million deaths (1) (March 2021). Most of the patients infected by SARS-CoV-2 are asymptomatic or present mild-moderate symptoms. However, in our midst, approximately 10% of patients require hospital admission (2) and 20% of hospitalized patients develop severe respiratory diseases which may quickly progress to respiratory failure, shock and multiorgan dysfunction (3). In hospitalized COVID-19 patients, more than 30% may require intensive care treatment and around 39% of ICU admitted patients die (4). Thus, it is essential to identify those individuals more likely to develop severe forms of the disease for an early precision management. Older age, male sex and pre-existing conditions such as hypertension, obesity, diabetes and chronic kidney disease are risk factors predisposing to serious disease. However, many individuals with these features do not develop severe symptoms, and the causes are not fully understood (3, 5).

The renin-angiotensin-aldosterone system (RAAS), one of the major blood pressure regulatory pathways, could be involved in the pathogenesis of COVID-19, since SARS-CoV-2 uses angiotensin-converting enzyme-2 (ACE2) as the binding receptor to enter the cell. The physiological homeostasis of this system is regulated by the balance of the angiotensin converting enzyme (ACE) and ACE2. ACE converts angiotensin I (Ang I) to angiotensin II (Ang II) which, through its interaction with the angiotensin II type 1 receptor (AT1R), induces a strong vasoconstriction and triggers proinflammatory, proapoptotic and profibrotic pathways in the lung and other organs (6). In RAAS, ACE2 contributes to the inactivation of Ang II, by hydrolyzing it to Ang-1-7, and, therefore, physiologically counters Ang II/AT1R effects, at the same time that stimulates vasodilation and promotes anti-inflammatory, antifibrotic and antithrombotic actions via the Ang-1-7/Mas receptor axis (6). In conditions where ACE 2 bioavailability is decreased, Ang II-AT1R pathway is potentiated, aggravating COVID-19-induced inflammation and lung injury. Also, high levels and activity of ACE would further increase the activity of the AngII/AT1R pathway.

Genetic polymorphisms in ACE, such as rs4343, rs4341 and the ACE I/D polymorphism, has been shown to affect ACE levels and activity, and confers susceptibility to hypertension (7), type 2 diabetes (8), overweight (9), nephropathy (10, 11) and certain cardiovascular (12, 13) and autoimmune diseases (14). Therefore, variants in ACE could also explain the different susceptibility to COVID-19. Indeed, an association of the DD genotype of the ACE I/D polymorphism with severe COVID-19 has been reported in hypertensive males (15). However, analyzing the I/D polymorphism is laborious and time-consuming and some authors have described a preferential amplification of the D allele (16). Rs4341 and rs4343 polymorphisms are in complete linkage disequilibrium with the ACE I/D polymorphism (16) and can be easily evaluated by TaqMan assays, therefore they could be better prognostic markers. In the present study we investigated the association of rs4341 and rs4343 polymorphisms of the ACE gene with COVID-19 outcomes in patients with different degree of severity.

## MATERIAL AND METHODS

### Subjects

In this study, 128 SARS-CoV-2 positive patients (confirmed by PCR from nasopharyngeal swabs) from the first epidemic wave (17 April −29 May 2020), were recruited at the Hospital San Pedro, Logroño, Spain. These patients were grouped into the following severity of illness categories: i) asymptomatic or mildly symptomatic patients not requiring hospital admission (n=33), ii) severe COVID-19 patients requiring hospitalization in the normal ward (n=66) and iii) critically ill patients admitted to the intensive care unit (ICU) (n=29). Relevant clinical data were obtained from the medical records of patients. The body mass index (BMI) of the patients was calculated as kg/m^2^. Patients were classified by BMI according to the criteria of the World Health Organization (WHO) and the Spanish Society for the Study of Obesity (SEEDO), with obesity defined as a BMI of 30 kg/m^2^ or higher for both sexes. This study was performed following the Helsinki Declaration and was approved by an independent ethical committee for clinical research (*Comité de Ética de Investigación con medicamentos de La Rioja*, CEImLAR, reference number PI-412). All patients or their representatives/relatives gave their consent to participate in the study.

### Genomic DNA isolation and genotyping

Genomic DNA was extracted from buffy coat layer using the DNeasy blood and tissue kit (Qiagen, Hilden, Germany). The rs4341 and rs4343 genotyping of the DNA samples was carried out using predesigned TaqMan SNP Genotyping Human Assays from Applied Biosciences (Foster City, CA) according to the manufacturer’s instructions.

### Statistical analysis

Categorical data are presented as counts (percentages) and were analyzed using the χ^2^ test or Fisher’s exact test. Quantitative values are expressed as mean ± standard deviation. Normal distribution of quantitative variables was checked using the Shapiro-Wilk test. Quantitative data were evaluated with the Kruskal-Wallis test followed by the Mann-Whitney U test. *P* values <0.05 were considered statistically significant. Statistical analysis was performed using GraphPad Prism 6 (GraphPad Prism®, La Jolla, California, USA).

Allele and genotype frequencies, Hardy-Weinberg equilibrium test, and association of the rs4341 and rs4343 polymorphisms with COVID-19 outcome were analyzed using SNPStats online software (https://snpstats.net/) (17). To reduce the potential confounding effects from age and gender, an adjustment for these factors was performed.

## RESULTS

### Distribution of demographic and clinical characteristics

Table 1 describes the characteristics and demographic data of SARS-CoV-2 infected patients. Mean age of asymptomatic/mildly symptomatic patients was lower than those with more serious illness (p<0.001). A higher frequency of males was found in both groups of hospitalized patients when compared to asymptomatic/mildly symptomatic patients (p<0.01 Hospital ward *vs* asymptomatic/mildly symptomatic; p<0.001 ICU *vs* asymptomatic/mildly symptomatic). A higher frequency of hypertension (p < 0.001) and dyslipidemia (p < 0.001) was also found in both groups of hospitalized patients when compared to asymptomatic/mildly symptomatic patients, and a similar tendency was observed when examining the frequency of type 2 diabetes mellitus (p<0.01), heart conditions (p<0.05) and dementia or other neurological conditions (p<0.001), although the difference only reached statistical significance for hospitalized patients who did not required ICU admission. A higher proportion of patients with chronic kidney disease was also detected in ICU admitted group when compared to asymptomatic/mildly symptomatic group (p<0.05). Besides, we found a higher prevalence of obesity in the group of patients admitted to the ICU when compared to asymptomatic/mildly symptomatic patients group (p<0.01). The percentage of obese patients in the hospital ward group was also higher than in the asymptomatic/mildly symptomatic group, although the difference was not statistically significant. We did not observe significant differences in the prevalence chronic obstructive pulmonary disease (COPD) among the 3 groups studied. The mortality rate was higher in patients with severe symptoms compared to patients with mild or no symptoms who did not require hospitalization (p<0.05 Hospital ward *vs* asymptomatic/mildly symptomatic; p<0.001 ICU *vs* asymptomatic/mildly symptomatic). Besides, the number of patients who died within 90 days after COVID-19 onset was significantly superior in the ICU group than in the hospital ward group (p<0.01).

**Table 1.**
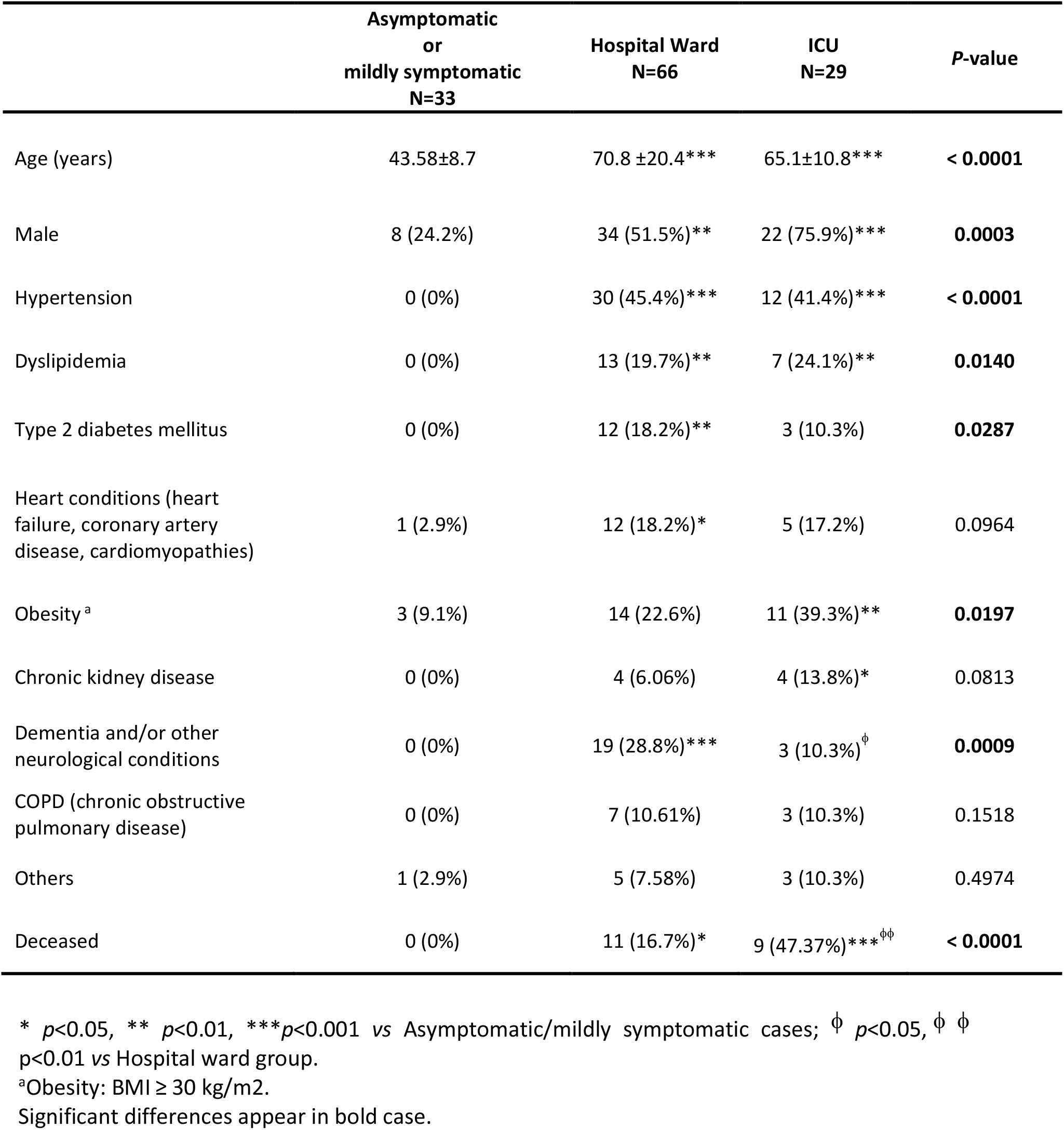
Clinical and demographic characteristics of COVID-19 patients.

### Genotype distribution among groups with different COVID-19 outcome

The genotypes of both polymorphisms were distributed in concordance with Hardy-Weinberg equilibrium in all groups (for rs4341: asymptomatic/mildly symptomatic *P*=0.16, hospital ward group *P*=1, ICU group *P*=0.72; for rs4343: asymptomatic/mildly symptomatic *P*=0.29, hospital ward group *P*=0.81, ICU group *P*=0.72).

We did not observe significant differences in the genotype and allele frequencies of rs4341 and rs4343 under any of the inheritance models between the 3 groups of SARS-Cov-2 infected patients (Figure 1). When we performed the analysis stratifying by sex no significant differences were observed when comparing rs4341 and rs4343 genotypes of male and female patients (Figure 2). When evaluating the association of both polymorphisms with the age of the patients and its interaction with the COVID-19 outcome no significant differences were observed either (data not showed).

**Figure 1.**
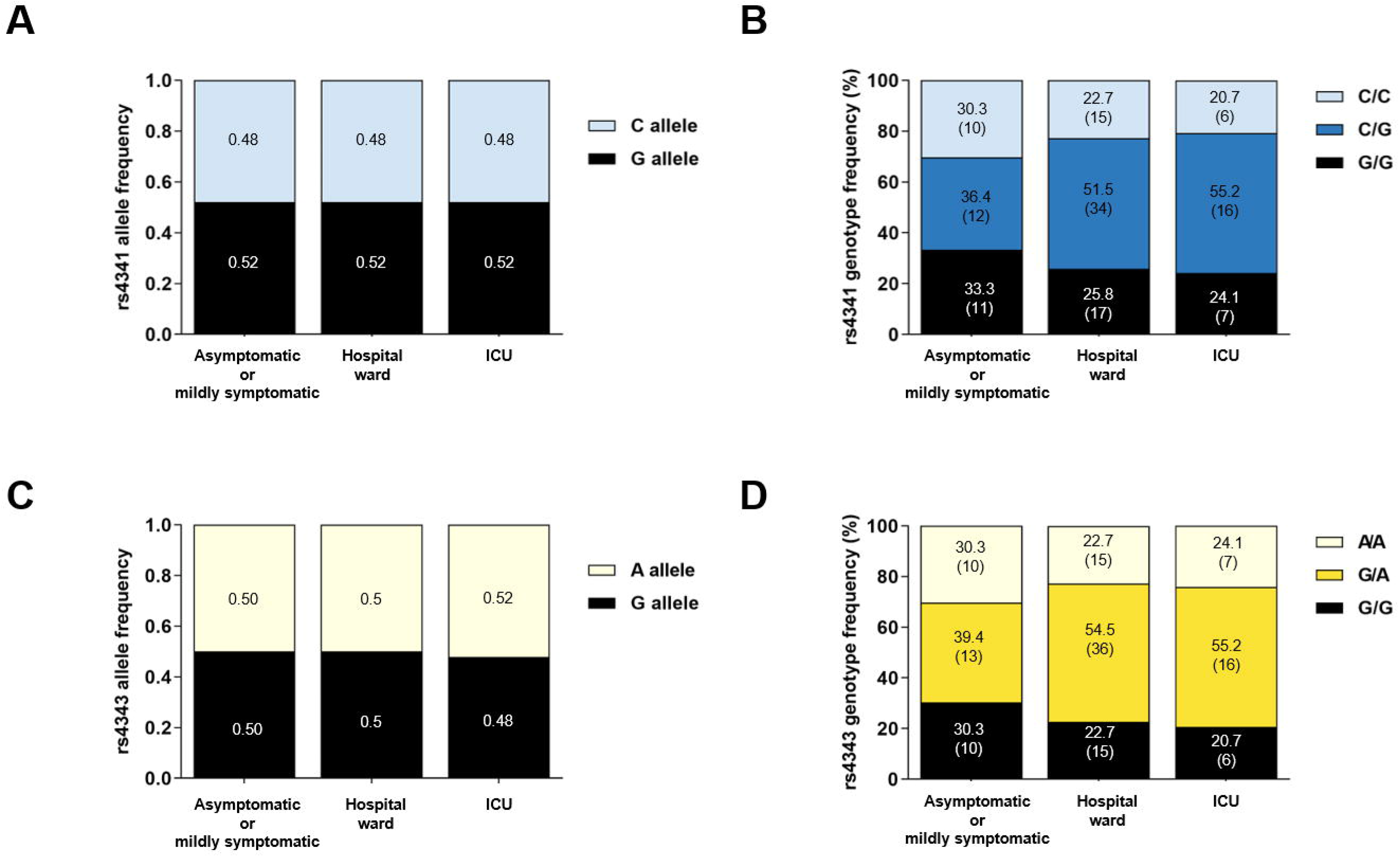
Allele and genotype distribution of the rs4341 and rs4343 polymorphisms among SARS-CoV-2 infected patients. Allele frequencies of rs4341 (A) and rs4343 (C) polymorphisms. Data are presented as proportions. Genotype frequencies of rs4341 (B) and rs4343 (D) polymorphisms. Data are presented as percentages. The number of patients is indicated in brackets.All data were adjusted for age and sex.

**Figure 2.**
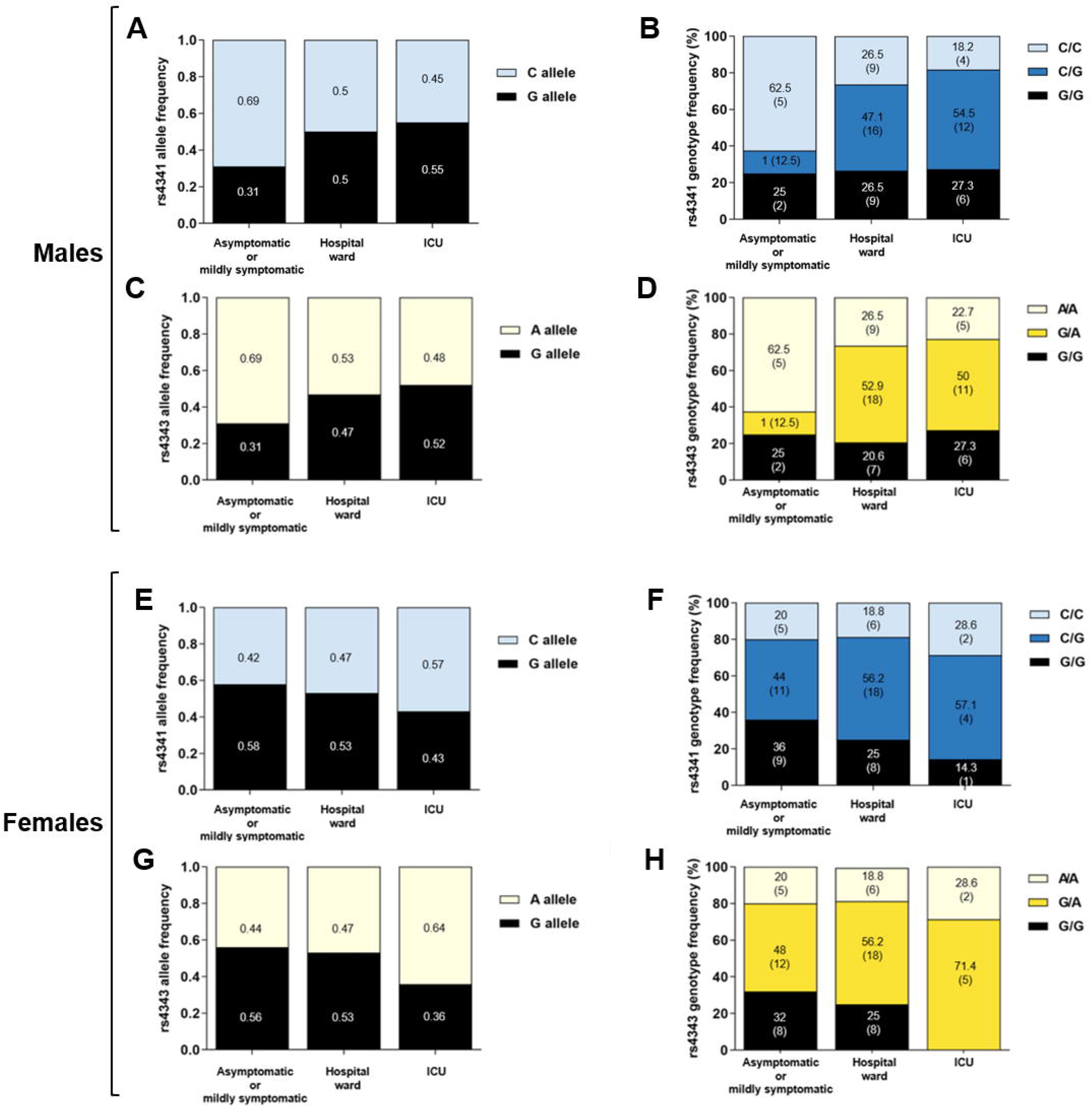
Allele and genotype distribution of the rs4341 and rs4343 polymorphisms among SARS-CoV-2 infected male and female. Allele frequencies of rs4343 and rs4341 polymorphisms among SARS-CoV-2 infected males (A and C) and females (E and G). Data are presented as proportions. Genotype frequencies of rs4343 and rs4341 polymorphisms among SARS-CoV-2 infected males (B and D) and females (F and H). Data are presented as percentages. The number of patients is indicated in brackets.All data were adjusted for age.

### Genotype distribution among groups with different COVID-19 outcome in relation to comorbidities

The association of the rs4341 and rs4343 polymorphisms with the risk of severe COVID-19 was also evaluated in relation to comorbidities. When we analysed only hypertensive patients admitted to the ICU, a higher frequency of the G allele in both polymorphisms compared to hypertensive patients hospitalized in the ward was observed (p<0.05) (Figure 3A and C). A similar, but not statistically significant tendency was observed in the frequency of the GG genotype (Figure 3B and D). When examining the genotypes of dyslipidemic patients divided according to COVID-19 severity, a higher frequency of G allele (p<0.01) and G-containing genotypes (p<0.05) was found in dyslipidemic patients admitted to the ICU than in those admitted to the hospital ward (Figure 3). The same differences were observed when evaluating the allele and genotype distribution of rs4341 and rs4343 in diabetic patients (p<0.001 for allele frequencies, p<0.01 for genotype frequencies) (Figure 3). These genotype frequencies observed in hypertensive, dyslipidemic and diabetic patients did not differ by sex (data not shown). A higher frequency of GG was not associated to a higher severity in obese patients with SARS-CoV-2 infection (data not shown). No associations were also found among rs4341 and rs4343 genotypes and COVID-19 severity in relation to heart conditions, renal disease, COPD or dementia and/or other neurological conditions (data not shown).

**Figure 3.**
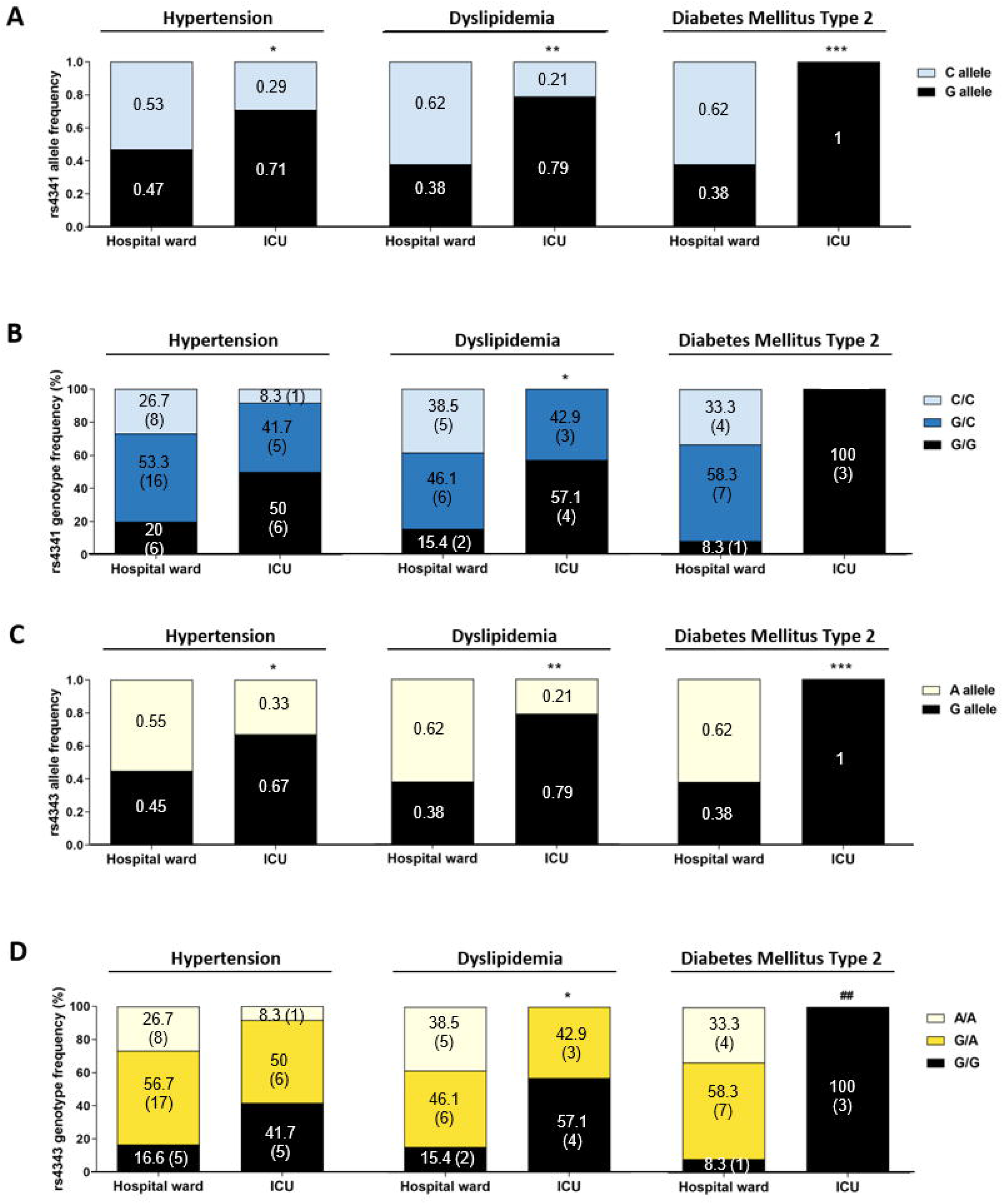
Allelic and genotypic frequencies of rs4341 and rs4343 polymorphisms in COVID-19 patients according to its hypertensive, dyslipidemic or diabetic status. Allele frequencies of rs4341 (A) and rs4343 (C) polymorphisms. Data are presented as proportions. * *p*<0.05, \*\**p*<0.01, *** *p*<0.001 *vs* Hospital ward. Genotype frequencies of rs4341 (B) and rs4343 (D) polymorphisms. Data are presented as percentages. The number of patients is indicated in brackets. * *p*<0.05 *vs* Hospital ward group for G carriers (GG+GC *vs* CC and GG+GA *vs* AA). ^$$^ *p*<0.01 *vs* Hospital ward between GG and GC and CC, GG+GC and CC, GG and GC+CC.^##^ *p*<0.01 *vs* Hospital ward between GG and GA and AA, GG+GA and AA, GG and GA+AA. All data were adjusted for age and sex.

Finally, a higher incidence of G-containing genotypes of both polymorphisms was detected among deceased COVID-19 patients who had been admitted to the ICU compared to survivals (p<0.05) (Figure 4).

**Figure 4.**
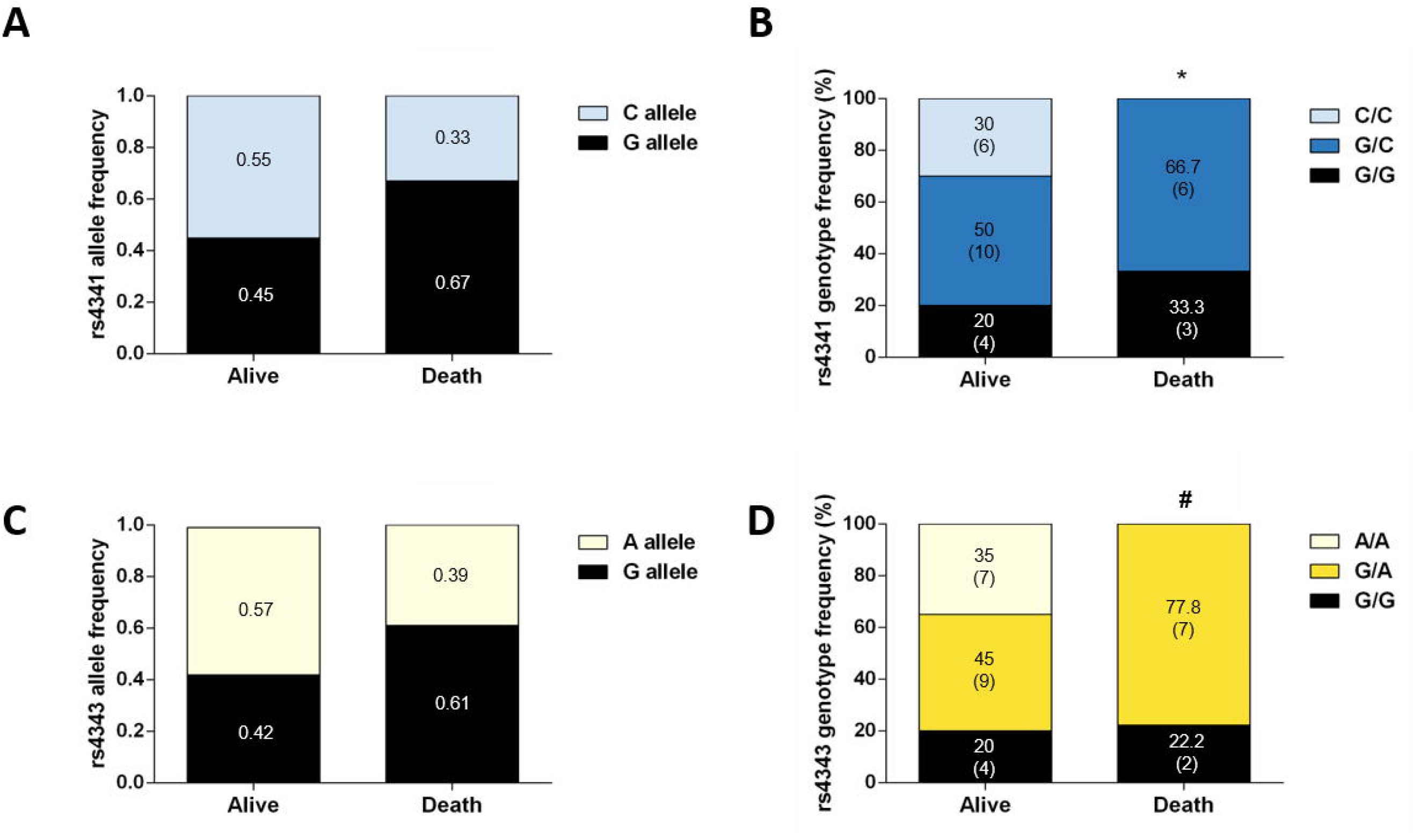
Genotype and allele frequencies of COVID-19 patients in ICU who survived or who died. Allele frequencies of rs4341 (A) and rs4343 (C) polymorphisms. Data are presented as proportions. Genotype frequencies of rs4341 (B) and rs4343 (D) polymorphisms. Data are presented as percentages. The number of patients is indicated in brackets.* *p*<0.05 *vs* survivors for G carriers (GG+GC *vs* CC). ^#^ *p*<0.05 *vs* survivors between GG and GA and AA, GG+GA *vs* AA).All data were adjusted for age and sex.

## DISCUSSION

Patients with SARS-CoV-2 infection can experience a wide range of clinical manifestations, ranging from no symptoms to critical illness and even death. However, the causes are still unknown. In this context, and although the sample size of the infected people is small, we found that COVID-19 severity was associated to male gender, as previously described (18). Similarly, older age and a higher number of comorbidities was found in both hospitalized groups compared to asymptomatic patients, in concordance with the literature (3, 19). However, we did not observe a higher mean age or a higher prevalence of hypertension, dyslipidemia, diabetes mellitus, obesity and cardiovascular disease in patients admitted to the ICU compared to those admitted to the ward, suggesting that other mechanisms/pathways could be involved in the severity of COVID-19 illness. In fact, several studies have suggested that variabilities in the genotype distribution of ACE polymorphisms could explain the variable prevalence and clinical outcomes of COVID-19 among different regions of the world (20-22). Thus, we investigated the association of two ACE polymorphisms involved in diabetes and hypertension (rs4341 and rs4343) (7, 8) with COVID-19 outcome in patients from La Rioja. In our study, we did not find differences for rs4341 and rs4343 frequencies among the three groups of patients analyzed when it was assessed without considering the comorbidities. However, we observed that having G allele in rs4341 and rs4343 worsens the outcome of hypertensive, dyslipidemic, and diabetic COVID-19 patients. Moreover, we found that, unlike hospitalized patients in the ward, all diabetic patients admitted to the ICU had the GG genotype. The number of type 2 diabetic patients in this group was very small (only 3 patients), so this result should be viewed with caution. However, our results clearly revealed that rs4341 and rs4343 polymorphisms of the ACE gene are related to the risk of developing severe COVID-19 (ICU admission) in hypertense, dyslipidemic and diabetic patients.

Interestingly, we further found a higher prevalence of the GG, GC and GA genotypes (all containing the G allele) among deceased UCI-patients, confirming the deleterious effect of the rs4341 and rs4343 G allele in COVID-19 outcomes. Precisely, 66% of these patients were hypertensive, thus, the presence of this polymorphism could be involved in the hypertension process *per se* as previously mentioned but also to be a synergetic risk factor for COVID-19 fatality. Some studies have reported an association of the GG genotype of rs4343 polymorphism and higher circulating levels and activity of ACE (23, 24), which could explain the higher susceptibility to develop severe forms of the disease in patients with the GG genotype, in addition to hypertension and dyslipidemia. SARS-CoV-2 sequesters ACE2 to invade cells, decreasing the bioavailability of ACE2 which entails a reduction in the degradation of Ang II and an exacerbation of the damaging effects of Ang II (25). Thus, the lung injury and inflammation caused by the reduced ACE2 levels due to the viral infection, and also by the hypertension, the dyslipidemia and diabetes, may be worsened by ACE genotypes that further increase ACE levels, and hence Ang II levels, such as the GG genotype of rs4341 and rs4343 herein analysed.

There are obviously several limitations in our study, such as the small size of our cohort. Thus, studies with higher number of patients could be of interest to clarify these results. Large-scale GWAS (Genome Wide Association Study) combined with WGS (whole genome sequencing) studies, such as the one being carried out by the SCOURGE consortium(26), will be very interesting to corroborate these results and to explore if they are extrapolable to the whole Spanish cohort or, in contrast, it is specific to our region.

In conclusion, the G-containing genotypes of rs4341 and rs4343 confer an additional risk factor of developing severe forms of COVID-19 in patients with hypertension, dyslipidemia, or possibly with diabetes independently of gender. These genotypes are also associated with an increased risk of death, mainly in hypertensive patients. Thus, the genotyping of rs4341 and rs4343 in COVID-19 patients could facilitate a more appropriate clinical management at admission.

## Data Availability

The datasets generated during and/or analysed during the current study are available from the corresponding author on reasonable request.

## ABBREVIATIONS

COVID-19: (coronavirus disease 19)
SARS-CoV-2: (severe acute respiratory syndrome coronavirus 2)
ACE: (angiotensin converting enzyme)
RAAS: (renin-aldosterone-angiotensin system)
ICU: (intensive care unit)
ACE2: (angiotensin-converting enzyme-2)
Ang I: (angiotensin I)
Ang II: (angiotensin II)
AT1R: (angiotensin II type 1 receptor)

## ACKNOWLEDGMENTS

We would like to thank the physicians and staff from the Infectious Diseases Department and from the Intensive Medicine Unit (Hospital San Pedro) for helping us with patient and sample recruitment. We would also like to thank Jorge Lázaro Galán and Dra. Lara García Álvarez for helping us with the collection of demographic and clinical data. We would also like to thank all patients and individuals who volunteered to participate in this study.

## AUTHOR CONTRIBUTIONS

MI, PP, and JAO designed the study, interpreted the data and wrote the manuscript. MI, PVB, ERF, and PPM performed the experiments. DEP, JA and MLF participated in patient and sample recruitment. All authors reviewed and/or edited the manuscript, and approved the submitted version.

